# Statistical Analysis Plan for the Blood Pressure Postpartum (BP^2^) study

**DOI:** 10.1101/2023.04.25.23289082

**Authors:** Bronwyn K Brew, Georgina M Chambers, Damian Kotevski, Amanda Henry, the BP2 Steering Committee

## Abstract

**Background:** Hypertensive Disorders of Pregnancy (HDP) complicate 5-10% of pregnancies globally: 2-5% preeclampsia, 3% gestational hypertension, and 0.5-2% chronic hypertension. It is epidemiologically well established that adverse health effects of HDP do not end with the pregnancy: these conditions are associated with increased lifetime cardiovascular disease (CVD) risks. However, intervention trial data in the early years postpartum after HDP, focussed on early intervention to reduce women’s ongoing CVD risks after HDP, is sparse.

**Methods:** The Blood Pressure Postpartum Trial (BP^2^) is a 3-arm randomised controlled trial investigating three different methods of follow-up and lifestyle behaviour change in the first 12 months postpartum following a pregnancy affected by HDP. The trial was prospectively registered on the Australian and New Zealand Clinical trials registry (registration: ACTRN12618002004246) and the study protocol has previously been published (Henry A et al, Pregnancy Hypertension 2020; 22: 1-6). This manuscript details the pre-analysis, pre-specified Statistical Analysis Plan.

## 2 Introduction

### 2.1 Preface

Hypertensive Disorders of Pregnancy (HDP) complicate 5–10% of pregnancies globally: 2–5% preeclampsia (PE), 3% gestational hypertension (GH), and 0.5–2% chronic hypertension (CH). It is well established that HDP’s adverse health effects do not end with the pregnancy: these conditions are associated with increased lifetime cardiovascular disease (CVD) risks. After HDP, women not only have a 3–4 times increased risk of chronic hypertension within 15 years, but a 2–3 times increased risk of CVD. This relationship is strongest for PE and CH but also exists for GH. Intervention data specific to the HDP population remain scarce. There has not yet been a randomised trial of specialised follow-up after HDP versus usual care, nor a sufficiently powered RCT after HDP to assess effect of lifestyle behaviour change strategies on presumed markers of later cardiovascular risk such as blood pressure and weight.

### 2.2 Scope of the analyses

The Blood Pressure Postpartum study (BP^2^) will provide evidence regarding the feasibility and effectiveness of postpartum lifestyle behaviour change interventions and structured clinical follow-up in improving cardiovascular health markers after HDP. This SAP applies to the 12-month follow up.

## 3 Study Objectives and Endpoints

### 3.1 Study Objectives

**In women who have had a hypertensive disorder of pregnancy,** to compare the effect of three postpartum (PP) managementstrategies at 1 year PP, withfurther observational follow-up at 3 years PP (note that in this SAP we are referring to the 1 year/12 month follow from now on). We willcompare the effects of:

1) **OptimisedUsualCare:** Standard postpartum care afterHDP,with the additionof information packages sent to both womenand their primaryhealthcare practitionersprior to a 6-months postpartum visit with the primary healthcare practitioner
2) **Brief Education Intervention:** One-off cardiovascular risk assessment and diet/exercise education in a dedicated clinic 6 months postpartum
3) **Extended Lifestyle Intervention:** Cardiovascular risk assessment and diet/exercise education in a dedicated clinic 6 months postpartum, followed by an individualised dietary and exercise lifestyle program of six months duration (from 6-12 months PP)

on

- *Maternal blood pressure* (BP)
- *Maternal positive lifestyle behaviour change*
- *Other maternal clinical cardiovascular risk indicators*
- *Non-invasivematernal measures of vascular structure and function*
- *Women’s health-related quality of life measures*
- *Infant growth and weight trajectories*
- *Healthcare costs, taking into account projected long-term savings*

### 3.2 Endpoints

#### Hypotheses

<u>One year postpartum, the Extended Lifestyle Intervention will, compared to Brief Education Intervention and Usual Care:</u>

1. significantly reduce maternal BP
2. result in superior maternal positive lifestyle behaviour change (as measured by BMI, waist circumference, physical activity and vegetable and fruit intake)
3. improve maternal clinical CVD risk indicators including lipid profile, cardiovascular risk score, HOMA [insulin resistance] score
4. improve maternal vascular structure and function as measured through non-invasive central arterial pressure, velocity and stiffness measures (with additional pilot subgroup investigation of utility of brachial Flow-Mediated dilatation, carotid Intima-Medial Thickness, hsCRP and other serum inflammatory markers). Note: COVID-19 pandemic and suspension of these measures means sample-size underpowered to test this hypothesis, vascular structure and function will be analysed as a separate substudy/exploratory data.
5. result in improved maternal health-related quality of life
6. demonstrate cost-effectiveness when long-term healthcare costs are considered
7. result in a more desirable infant growth trajectory and adiposity (by weight, length and BMI centiles)

Additional hypothesis: The effects observed in Hypotheses 1-7 will continue to be observed at 3 years postpartum for mothers and infants.

## 4 Study Methods

### 4.1 General Study Design and Plan

(ICH E3;9)

<u>Study configuration and experimental design</u>: three-arm, multicentre, randomized controlled trial with participants allocated 1:1:1 to control group, Brief Education Intervention and Extended Lifestyle Intervention.

PROBE design: Prospective Randomized Open Blinded Endpoint.

<u>Type of Comparison</u>: Superiority of intervention groups versus control

<u>Type of control</u>: optimized usual care

<u>Level and method of blinding</u>: Open blind, patients and clinicians will not be blinded, however clinicians taking outcome measurements and analyst team will be blinded to group allocation.

<u>Method of treatment assignment</u>: Randomization with stratification by study hospital, parity and body mass index (see Section 4.3).

### 4.2 Inclusion-Exclusion Criteria and General Study Population

(ICH E3;9.3. ICH E9;2.2.1)

**General Study population**: Australian teaching hospitals providing maternity care where staffing includes specialist obstetricians, physicians, paediatricians and anaesthetists/critical care specialists and >1500 births per year.

**Inclusion criteria:**

- Aged ≥18 years
- Given birth at one of the study hospitals (St George Public Hospital, Royal Prince Alfred Hospital, Liverpool Hospital, Campbelltown hospital, Royal Hospital for Women, Westmead Hospital) during the study period
- The index pregnancy was complicated by a hypertensive disorder of pregnancy (HDP): preeclampsia (PE), gestational hypertension (GH), chronic hypertension (CH), chronic hypertension plus superimposed preeclampsia (CH+PE)

**Exclusion criteria:**

- Stillbirth or neonatal death from HDP pregnancy
- Planning to move out of study area/unavailable for follow up during study period
- Active severe mental health condition or developmental disability precluding informed consent.

**Participant Flow Diagram**

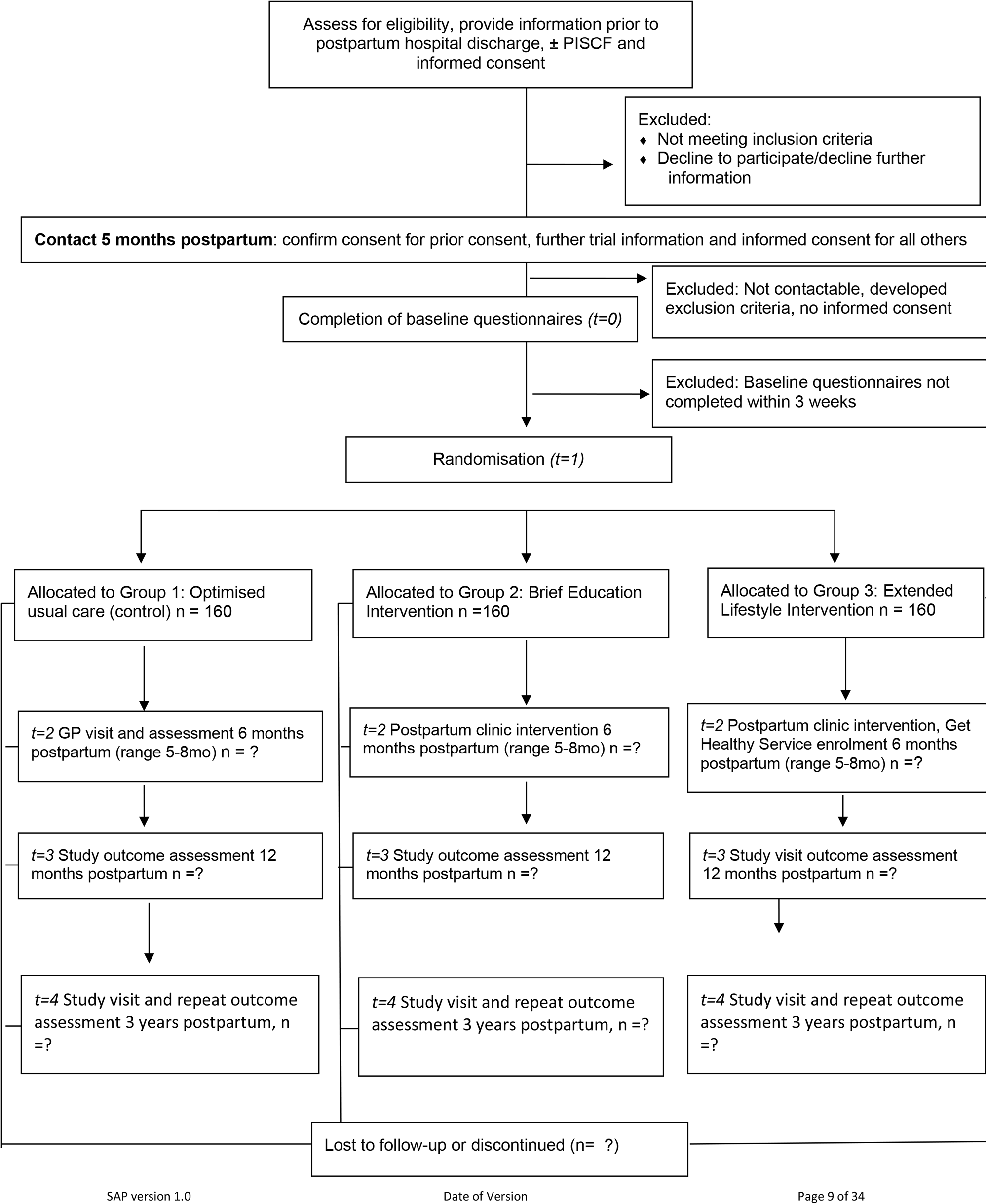

### 4.3 Randomization and Blinding

(ICH E3; 9.4.3, 9.4.6. ICH E9; 2.3.1, 2.3.2)

Participants will be randomly assigned to Group 1, 2 or 3 with a 1:1:1 allocation as per a computer-generated randomisation schedule using REDCap software. Randomisation will be stratified by site, parity (1 versus >1) and body mass index (<=30 versus 30+), using blocks. Details relating to block randomisation will be kept in a separate document not available to anyone who is involved in trial recruitment. Block size will be revealed after analysis. Allocation concealment is ensured, as the randomisation code will not be released until after baseline questionnaires have been completed and the study researcher then accesses REDCap for randomisation to Group 1, 2 or 3.

All participants who give consent for participation and who complete the baseline questionnaires within the scheduled 3 week timeframe will be randomised. Randomisation will be performed in REDCap by staff responsible at each study site for recruitment, which will usually be the study nurse/midwife at that site. This staff member will then organise information package and instructions regarding GP follow-up (Group 1) or for information package and postpartum clinic visit (Groups 2 and 3). This staff member will not be involved in outcome assessment in postpartum clinic at 6 month, 12 month or subsequent visits.

Due to the nature of the intervention, neither postpartum clinic staff nor participants can be blinded to allocation. Regarding analysis, a PROBE design (prospective randomised open blinded endpoint) is adopted, with analysis occurring blinded to group allocation (Ford & Norrie, 2016). After all participant data including outcome data is entered by study staff, it will be exported with group allocation concealed for blinded analysis by the trial statistician and trial health economist as applicable.

### 4.4 Study Assessments

(ICH E3; 9.5.1. ICH E9; 2.2.2)

#### 4.4.1 Timing of assessments

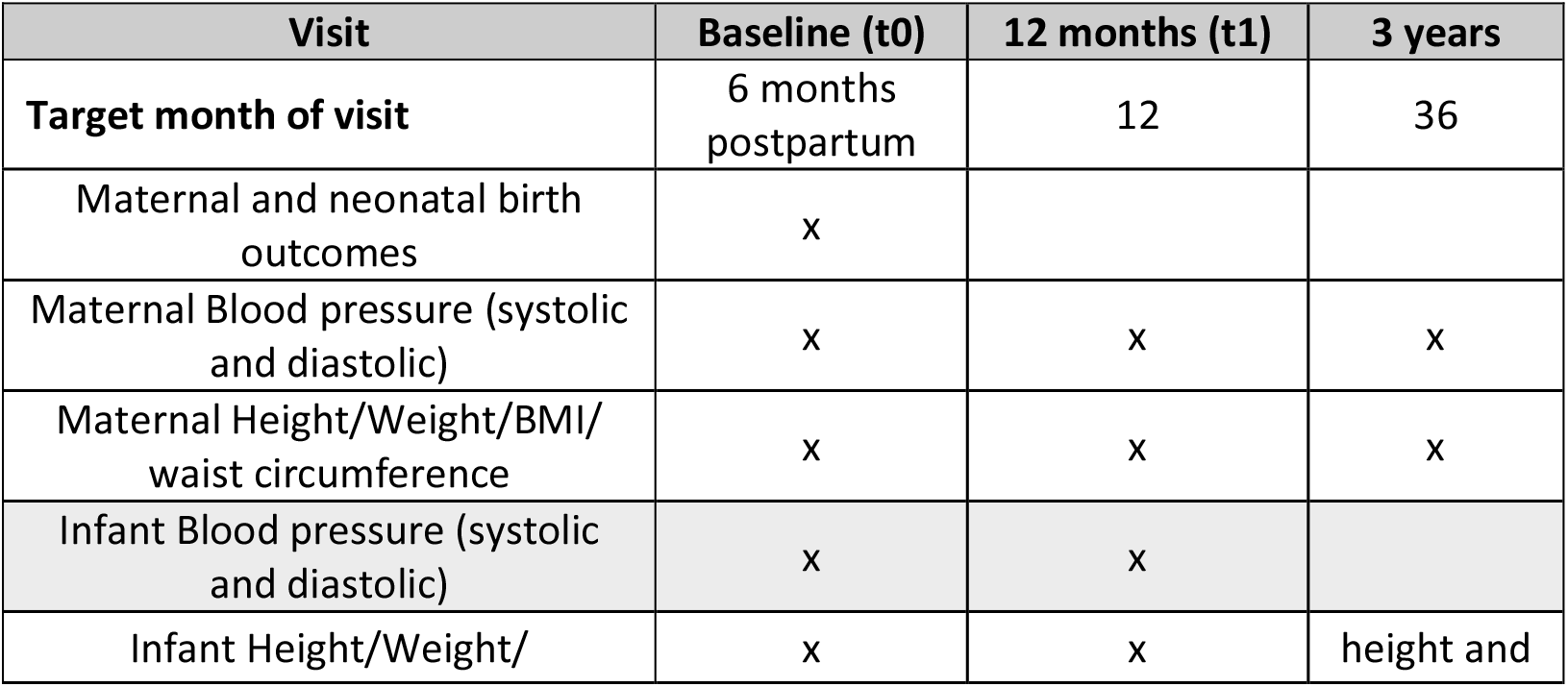
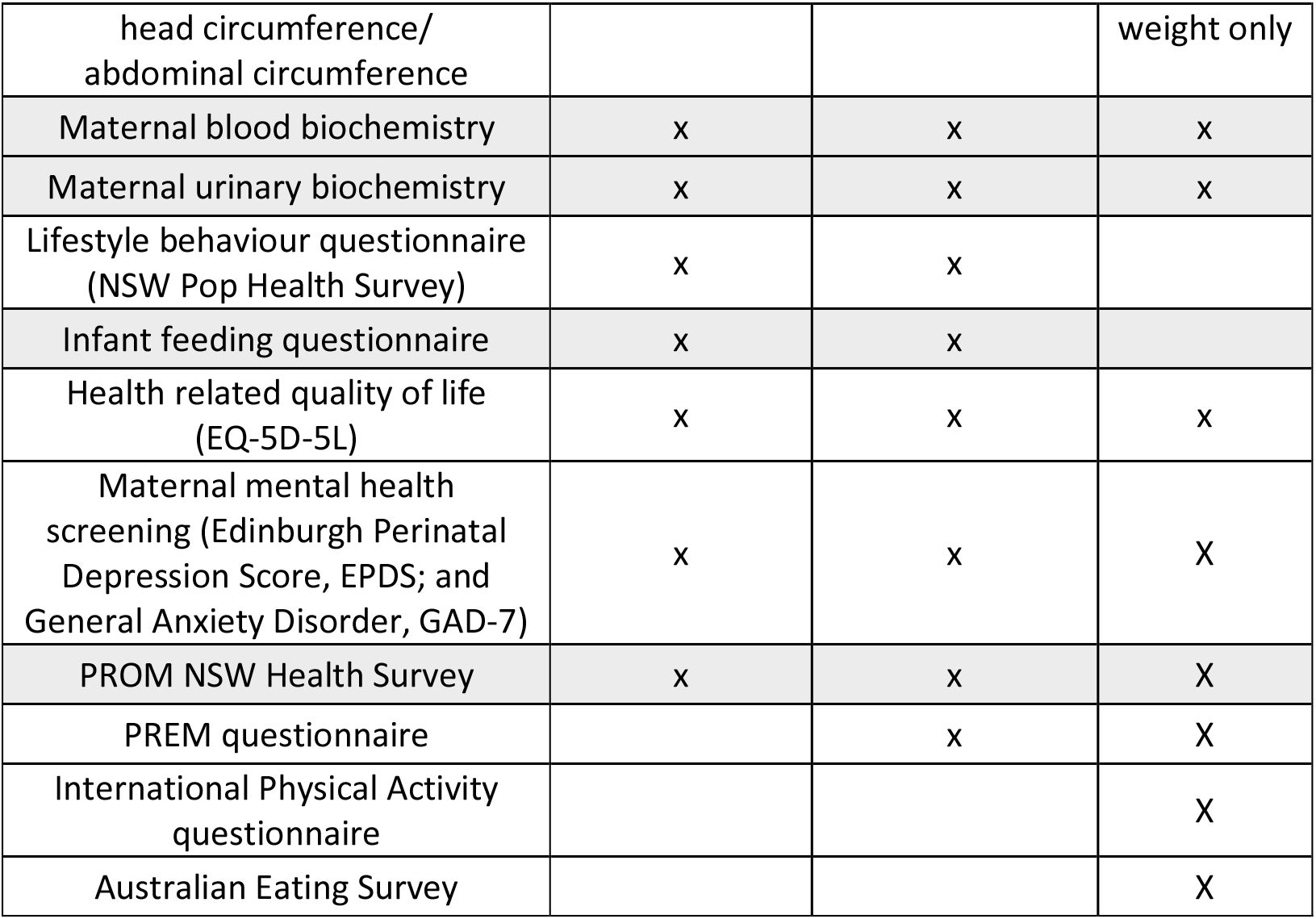

#### 4.4.2 Analysis time windows

| Visit (target month) | Lower bound | Upper bound |
| --- | --- | --- |
| Baseline (6 months) | 5 months | 8 months |
| 12 months | 12 months | 15 months |
| 36 months | 36 months | 48 months |

To allow flexibility for this pragmatic trial during the COVID-19 pandemic, trial entry was relaxed from upper bound of 7 months postpartum to 8 months postpartum, and collection of 12-month outcome data upper bound of 14 months to 15 months. Time between entry and collection of primary outcome data to be minimum of 5 months. Data outside the time windows is not accepted for use in analysis.

#### 4.4.3 Variables

**Variable measurements**

| Variable | Type of Variable | Range/categories |
| --- | --- | --- |
| **Descriptives** | (for Table 1) |  |
| Maternal age | Continuous<br>(from hospital info) |  |
| Maternal Country of Birth | Categorical (baseline<br>information) | 1= Australia<br>2 =New Zealand<br>3 =Asia<br>4 =Europe and United<br>Kingdom |
|  |  | 5 =Middle East/North Africa<br>6 =Africa<br>7 =North America<br>8 =South America<br>9 =Other |
| Maternal ethnicity | categorical<br>(Baseline questionnaire) | 1 Caucasian<br>2 Asian<br>3 Aboriginal or Torres Strait Islander<br>4 Polynesian<br>5 European<br>6 Middle Eastern<br>7 African<br>8 other |
| Maternal highest Education level | Categorical<br>(baseline questionnaire) | 1 Secondary School<br>2 Trade/Cert/Diploma<br>3 University |
| Gravidity | Categorical (hospital info) | 1<br>2 or more |
| Parity | Categorical (hospital info)<br>Categorical | 1<br>2<br>3 or more |
| Plurality | Categorical (hospital info) | 1<br>2 (twins) |
| Smoker during pregnancy | Binomial<br>(hospital info) | 0 no<br>1 yes |
| Current Smoker at 6 months | Binomial<br>(lifestyle questionnaire or Postpartum clinic questionnaire) | 0 no<br>1 yes |
| Pregnancy systolic blood pressure at booking | Continuous (hospital info)<br>Ordinal | Continuous mmHg<br>1= <120mmHg<br>2= 120-129mmHg<br>3= 130-139mmHg<br>4 = 140mmHg or above |
| Pregnancy diastolic blood pressure at booking | Continuous (hospital info)<br>Ordinal | Continuous mmHg<br>1 = <80mmHg<br>2= 80-84 mmHg<br>3= 85-89mmHg<br>4= 90mmHg or above |
| Systolic Blood pressure at baseline (t0) | Continuous<br>Ordinal | Continuous mmHg<br>1= <120mmHg<br>2= 120-129mmHg<br>3= 130-139mmHg |
|  |  | 4 = 140mmHg or above |
| Diastolic Blood pressure at baseline (t0) | Continuous<br>(hospital info)<br>Ordinal | Continuous mmHg<br>1 = <80mmHg<br>2= 80-84 mmHg<br>3= 85-89mmHg<br>4= 90mmHg or above |
| Weight at baseline (t0) | Continuous (kg) |  |
| Waist circumference at baseline (t0) | Continuous (cm) |  |
| BMI at baseline (t0) | 1) Categorical | 1 <30 kg/m<sup>2</sup><br>2 ≥ 30 kg/m<sup>2</sup><br>Extended categorical:<br>1 Underweight (BMI <18.5 kg/m<sup>2</sup>)<br>2 Normal weight (BMI 18.5-24.9 kg/m<sup>2</sup>)<br>3 Overweight (BMI 25.0-29.9 kg/m<sup>2</sup>)<br>4 Obesity Class I-II (BMI 30.0-39.9 kg/m<sup>2</sup>)<br>5 Obesity Class III/severe obesity (BMI 40 kg/m<sup>2</sup> or greater) |
|  | 2) Continuous | BMI in kg/m<sup>2</sup> |
| Type of hypertension during pregnancy | Categorical<br>(hospital info) | 1 chronic hypertension<br>2 gestational hypertension<br>3 preeclampsia<br>4 preeclampsia and chronic hypertension |
| Other pregnancy complications during the index pregnancy (gestational diabetes, Cervical suture, cholestasis, antepartum haemorrhage, fetal growth restriction, preterm premature rupture of members, threatened premature labour, spontaneous preterm labour, other) | Binomial<br>(hospital info) | 0 no<br>1 yes |
| Gestational diabetes (GDM) | Binomial (hospital info) | 0 no<br>1 yes |
| Obstetric and fetal Complications (threatened preterm labour, antepartum haemorrhage, fetal, growth restriction) | Binomial (hospital info) | 0 no<br>1 yes |
| Other Medical complications of pregnancy (cholestasis, pyelonephritis) | Binomial (hospital info) | 0 no<br>1 yes |
| History of hypertension, diabetes, renal disease or cardiac disease | Binomial<br>(hospital info and baseline questionnaire) | 0 no<br>1 yes |
| History of hypertension | Binomial<br>(hospital info and<br>baseline questionnaire) | 0 no<br>1 yes |
| History of diabetes (GDM, T1D, T2D) | Binomial<br>(hospital info and<br>baseline questionnaire) | 0 no<br>1 yes |
| History of renal disease | Binomial<br>(hospital info and<br>baseline questionnaire) | 0 no<br>1 yes |
| History of cardiac disease | Binomial<br>(hospital info and<br>baseline questionnaire) | 0 no<br>1 yes |
| Hypertensive Medication at baseline | Categorical | 0 no<br>1 yes |
| Mode(s) of Birth | Categorical (hospital<br>info) | 1= unassisted vaginal<br>birth<br>2= breech vaginal birth<br>3= assisted vaginal<br>birth (vacuum and/or<br>forceps)<br>4= elective caesarean<br>section<br>5= emergency<br>caesarean section |
| Infant sex | Binomial<br>(hospital info) | 1= male<br>2= female |
| Infant birth weight | Continuous (grams)<br>(hospital info) |  |
| Gestational age at birth | Continuous (weeks)<br>Ordinal | Continuous<br>1= Term (37+0 weeks<br>or more)<br>2= Late preterm (34+0-<br>36+6 weeks)<br>3= Moderately preterm<br>(32+0-33+6 weeks)<br>4= Very preterm (28+0-<br>31+6 weeks)<br>5= Extremely preterm<br>(<28+0 weeks) |
| Maternal Intensive Care or High<br>Dependency/Acute Care admission during<br>the birth admission | Binomial<br>(hospital info) | 0= no<br>1= yes |
| Infant neonatal intensive care or special care<br>nursery admission | Categorical (hospital<br>info) | 0= no<br>1 = yes |
| Current infant feeding at baseline (t0) | Binomial (baseline<br>questionnaire) | 1 = Breastfeeding<br>(exclusive or mixed)<br>0 = No breastfeeding<br>(Bottle/artificial feeding) |
|  |  | only) |
| **Primary outcomes** |  |  |
| Change in Systolic Blood pressure (12m -6m) | Continuous<br>Average of 3 readings to be used<br>Measured with blood pressure machine |  |
| Composite threshold primary outcome (12m -6m) | Binomial<br>(change in body weight or waist circumference) | 0 = change in body weight <4kg AND change in waist circumference <2cm<br>1= change in body weight ≥ 4kg AND/OR change in waist circumference ≥ 2cm |
| **Secondary Outcomes** |  |  |
| **Lifestyle Behaviour changes** |  |  |
| Self-rated general health baseline (t0) | Ordinal | 1 Excellent<br>2 Very Good<br>3 Good<br>4 Fair<br>5 Poor<br>6 Very Poor<br>X Don't know<br>R Refused |
| Self-rated general health 12m | Ordinal | As above |
| Self-rated mental health baseline (t0) | Ordinal | As above |
| Self-rated mental health 12m | Ordinal | As above |
| Self-rated wellbeing baseline (t0) | Ordinal | 1 Delightful<br>2 Pleased<br>3 Mostly satisfied<br>4 Mixed<br>5 Mostly dissatisfied<br>6 Unhappy<br>7 Terrible<br>X Don't know<br>R Refused |
| Self-rated wellbeing 12m | Ordinal | As above |
| Change in Alcohol Frequency days per month (12m-6m) | Continuous – number of days in a month<br>(Calculate from ALC1 questions on days/week, days/month) |  |
| Change in Average serves of vegetables per day (12m-6m) | Continuous- number of vegetable serves per day<br>(Calculate from NUT1 questions on vegetable serves each week and day) |  |
| Change in Average serves of fruit per day (12m-6m) | Continuous- number of fruit serves per day<br>(Calculate from NUT2 questions on vegetable serves each week and day) |  |
| Change in Average moderate-vigorous physical activity per week (12m-6m) | Continuous- number of hours per week<br>(Calculate adding PAC2/b (walking) + PAC4a/b (vigorous chores) + PAC 6a/b (gardening) + PAC 8a/b (vigorous physical activity) + PAC10a/b (moderate exercise) + PAC11a/b (muscle strengthening) |  |
| Change in Smoking status (12m-6m) | Binomial | 0= (never smoked, tried a few times but not regularly, previous smoker)<br>1= Smokes daily or occasionally |
| Following a healthy lifestyle at 12 months | Binomial<br>(12 month survey) | 1 yes<br>0 no |
| Confident that diet and exercise changes will be permanent | Ordinal<br>(12 month survey) | 1 not at all<br>2 slightly<br>3 somewhat<br>4 quite a bit<br>5 extremely |
| Signed up for any GHS by 12m | binomial | 0 = no<br>1= yes |
| **Maternal Clinical** |  |  |
| Hypertensive Medication (12m) | Categorical<br>(questionnaire 12m) | 0 No<br>1 Yes |
| Projected cardiovascular risk at 12m | Ordinal: number of risk factors above the reference range:<br>Systolic BP ($\geq 120$)<br>Diastolic BP ($\geq 80$)<br>BMI ($\geq 25.0$)<br>LDL cholesterol ($>4.0$)<br>Homeostatic assessment (HOMA) score ($> 3.0$)<br>Urinary albumin:creatinine ratio ($> 2.0$)<br>Current Smoker (yes) | 0 = none<br>1= 1 cardiovascular risk factor<br>2 = $>1$ cardiovascular risk factors |
| Change in Weight (12m-6m) | Continuous (kg)<br>Clinic/GP measures to be used when available<br>Self-report when clinic/GP values not available |  |
| Change in Diastolic BP (12m-6m) | Continuous |  |
| Systolic blood pressure 12m | Ordinal | 1= $<120$mmHg<br>2= 120-129mmHg<br>3= 130-139mmHg<br>4 = 140mmHg or above |
| Diastolic blood pressure 12m | Ordinal | 1 = $<80$mmHg<br>2= 80-84 mmHg<br>3= 85-89mmHg<br>4= 90mmHg or above |
| Change mean arterial pressure (MAP) 12m-6m | Continuous (mmHg) |  |
| Change in waist circumference (12m-6m) | Continuous (cm)<br>Measured following Get Healthy NSW online written instructions<br>Clinic/GP measures to be used when available<br>Self-report when clinic/GP values not available |  |
| Change in BMI (12m-6m) | Continuous |  |
| BMI category at 12m | Ordinal | 1 Underweight (BMI <18.5 kg/m<sup>2</sup>)<br>2 Normal weight (BMI 18.5-24.9 kg/m<sup>2</sup>)<br>3 Overweight (BMI 25.0-29.9 kg/m<sup>2</sup>)<br>4 Obesity Class I-II (BMI 30.0-39.9 kg/m<sup>2</sup>)<br>5 Obesity Class III/severe obesity (BMI 40 kg/m<sup>2</sup> or greater) |
| Change in waist:height ratio (12m-6m) | Continuous |  |
| CV risk baseline | Categorical<br>(calculated using Heart Foundation online calculator, uses gender, smoking status, age, systolic BP, total cholesterol, HDL cholesterol, diabetic status)<br>https://www.cvdcheck.org.au/ | 1= low risk<br>2= moderate risk (10-15% chance CVD in next 5 years)<br>3= high risk (>15% in next 5 years) |
| CV risk 12 months | As above | As above |
| **Maternal Biochemical** |  |  |
| Change in HOMA score (12m-6m) | Continuous |  |
| Change in fasting Insulin (12m-6m) | Continuous |  |
| Change in fasting glucose (12m-6m) | Continuous |  |
| Change in low density lipoprotein cholesterol (12m-6m) | Continuous |  |
| Change in high density lipoprotein cholesterol (12m-6m) | Continuous |  |
| Change in triglyceride cholesterol (12m-6m) | Continuous |  |
| Change in hsCRP (12m-6m) | Continuous |  |
| Change in urinary albumin: creatinine ratio (12m-6m) | Continuous |  |
| Change in serum creatinine (12m-6m) | Continuous |  |
| Change in eGFR (12m-6m) | Continuous |  |
| Change in AST Transaminase (12m-6m) | Continuous |  |
| Change in ALT Transaminase (12m-6m) | Continuous |  |
| Change in haemoglobin concentration (12m-6m) | Continuous |  |
| Change in platelet concentration (12m-6m) | Continuous |  |
| **Infant Measures** |  |  |
| Change in weight centile (12m-6m) | Continuous | Z scores based on WHO Child Growth Standards for infants $\geq 6$ months, and 21<sup>st</sup> Intergrowth Preterm Postnatal Growth Standards for infants $< 6$ months (corrected age) at 6 month visit |
| Infant feeding at 12m | Binomial | 1 = Breastfeeding (exclusive or mixed)<br>0 = No breastfeeding (Bottle/artificial feeding only) |

## 5 Sample Size

(ICH E3; 9.7.2. ICH E9; 3.5)

The proposed sample size is 480 women total (160 women in each arm). This sample size will power the trial (80% with two tailed, alpha of 0.05) to detect, for the co-primary outcomes, *either*

1) A difference of 15% in the proportion of women who achieve a 4 kg weight loss and/or 2 cm waist circumference reduction, from 70% in Group 1 to 85% in Group 2 or 3

OR

2) 4 mmHg difference in systolic BP between groups at 12 months postpartum (with baseline measures taken at 6 months postpartum). These calculations assume for Usual Care (Group 1), average weight 75 kg with standard deviation (SD) 19 kg, waist circumference 97 cm (SD 15 cm), and systolic BP 114 mmHg with SD 10 mmHg (values derived from our P4 cohort). Dropout rate is calculated at 3% from randomisation at 5.5 months to main outcome assessment at 12 months. This is based on loss to follow-up rate of only 4% between 6 months postpartum and 2 years postpartum for our existing P4 cohort study, and the high safety/low side effect potential of the interventions and the outcome assessment, meaning dropout and loss to follow-up in the short term is likely to be low.

## 6 General Analysis considerations

### 6.1 Timing of Analyses

The final analysis will be performed when:

- All randomised subjects have either completed the 12 month visit or dropped out of the study prior to 12 months.
- The data for analysis meets the cleaning and approval requirements of the main analyst:

- Attributing baseline levels to any missing 12 month data
- Checking missing data values for validity and consistency
- Checking data dictionary matches with dataset
- Removal of Group 3 GHS compliance questions (and any other questions that are only answered by one of the randomized groups)
- Removal of group names, randomisation of groups with numbers or letters so analyst is blinded
- Check for invalid entries eg birthweight of 1g and either fix or set to missing
- After the finalization and approval of this SAP document by the analyst, the principal investigator and the study coordinator.

### 6.2 Analysis Populations

(ICH E3; 9.7.1, 11.4.2.5. ICH E9; 5.2)

#### 6.2.1 Intention to Treat Population

##### Primary outcomes

All participants who were randomized at 6 months with baseline measures are included in the analysis. Primary outcome data for those lost to follow-up at 12 months will be handled by one of the following approaches:

1. Single imputation: participants will be assumed to have returned to baseline (6 months) and given their baseline measures for the primary outcomes.
2. Complete cases analysis: only participants with data at both baseline and 12 months will be included in the analysis.
3. Multiple imputation: a subset of the dataset will be used to impute the missing 12-month outcome data.

##### Secondary Outcomes

Full Analysis Set. Those with data at both baseline and 12 months for the outcome of interest. Number and missing to be reported for each outcome.

#### 6.2.2 Per Protocol Population

No per protocol analysis to be done.

### 6.3 Covariates and Subgroups

(ICH E3; 9.7.1, 11.4.2.1. ICH E9; 5.7)

Covariates: adjust analyses for hospital.

Subgroup analysis for the stratified randomisation and pre-specified groups (below).

Primary and secondary outcomes to be assessed (secondary results will go in supplement). Significance of the treatment*subgroup interaction term in the regression model at a level of p ≤ 0.01 will be required to conclude there exists heterogeneity of treatment effect.

- Parity (1 v >1)
- BMI (<30 v ≥ 30 at baseline)
- Type of hypertensive disorder of pregnancy (chronic hypertension, gestational hypertension, preeclampsia, preeclampsia superimposed on chronic hypertension)

#### 6.3.1 Multi-centre Studies

(ICH E3;9.7.1, 11.4.2.4. ICH E9; 3.2)

This study includes participants from 6 hospitals (St George, Royal Hospital for Women, Royal Prince Alfred, Liverpool, Westmead and Campbelltown). All analyses will be adjusted for hospital.

### 6.4 Missing Data

(ICH E3; 9.7.1, 11.4.2.2. ICH E9;5.3. EMA Guideline on Missing Data in Confirmatory Clinical Trials)

<u>End of recruitment</u>: discussion with study management team and project statistician to discuss the proportion of missing data and strategies to minimise missing data prior to final data cleaning and analysis.

Missing data will be assessed to determine whether the 12-month outcome data is missing at random. Strategies to deal with missingness will depend on the nature of missingness and could include:

1. Baseline data: impute a value of 0 for missing covariates, and then include a missing-indicator variable for the covariate as an additional covariate in the regression model [White and Thompson (2005)].
2. Outcome data: where GP, clinic-based measures are missing e.g. waist circumference, weight, then self-report measures can be used.
3. Lost to follow up: as specified with Intention to Treat, there are three possible approaches to handling the missing 12-month. Using baseline measures for primary outcomes in those who drop out is the best way to mimic real world practice where people do not continue with an intervention, however it has the potential to bias the results towards the null especially if there is a large loss to follow up, therefore, we will consider multiple imputation of missing data based on the nature of missingness.

Reporting: proportion of missing data at baseline and loss to follow up will be reported as well as methods to deal with both issues.

### 6.5 Multiple Testing

(ICH E3; 9.7.1, 11.4.2.5. ICH E9; 2.2.5)

The primary analysis compares 12 months (outcome, t=1) and 6 months (baseline, t=0), therefore only two points in time. Subsequent analyses at later times may include 12 month data and longitudinal analysis accounting for multiple time points, but that is not the focus of the current analysis.

However, in the initial analysis given that there are a number of primary and secondary outcomes this increases the risk of Type 1 error due to multiple hypothesis testing. This will be dealt with one of two methods: 1) by controlling for the false discovery rate (FDR) as proposed by Benjamini & Hochberg 1995 (J R Statistic Soc: 57; 289) or controlling for the familywise error rate (FWER) proposed by Hothorn, Bretz & Westfall 2008 (Biometric J:50; 346) through the R package multcomp. The Hothorn, Bretz & Westfall approach allows simultaneous inferences to be made over both parametric models and all relevant hypotheses, while accounting for correlation between the test statistics and allowing for stepwise testing procedures.

For the analysis of primary outcomes the type 1 error rate for either multiplicity testing method will be set to α= 0.05 and for the secondary outcomes α = 0.01.

## 7 Summary of Study Data

All continuous variables will be summarized using the following descriptive statistics: n, mean, standard deviation, median, maximum and minimum. The frequency and percentages of observed levels will be reported for all categorical measures. All summary tables will be structured with a column for each treatment in the order (Usual care, Brief Intervention, Extended Lifestyle Intervention, All participants) and will be annotated with the total population size relevant to that table/treatment, including any missing observations.

### 7.1 Subject Disposition

As shown in the Participant Flowchart above (page 9).

### 7.2 Derived variables

As shown in the 4.4.3 variable list.

### 7.3 Protocol Deviations (COVID-19 pandemic)

Due to the COVID-19 pandemic essential clinical and outcome measures (blood pressure, weight, and blood/urine tests) 6 and 12 months postpartum were performed, depending on LHD/site policy (a) by the research midwife, with visits kept to <15 minutes, and separate from antenatal/outpatient clinic spaces (i.e. no waiting time and woman able to maintain social distancing) (b) where possible via the woman’s GP, to coincide with necessary visits of her infant (for 6 and 12 month check and vaccinations).

Women were reminded that they and their baby must be well/asymptomatic of COVID-19 to be attending such visits. Therefore, there may be some missing data due to COVID-19 infections. These women will be treated as outlined in section 6.4 after consideration of the amount of missingness.

In addition, due to pandemic restrictions one of the secondary outcomes-maternal vascular structure and function was not measured. As this is a secondary outcome and the amount of missing data will be considerable this outcome will not be included in the primary analysis.

### 7.4 Demographic and Baseline Variables

As shown in Section 4.4.3 ‘Variables’ under ‘**Descriptives’**. Presented as outlined in Section 7 ‘Summary of study data’ and Table 1 Section 14.

### 7.5 Concurrent Illnesses and Medical Conditions

As shown in Section 4.4.3 ‘Variables’ under ‘**Descriptives’**. Presented as outlined in Section 7 ‘Summary of study data’ and Table 1 Section 14.

### 7.6 Compliance

For groups 1 and 2 all participants will be considered to have complied, as baseline (6 months visit) data cannot be obtained without participants having been compliant to this allocation. For group 3, participants will be considered (a) non-compliant if they did not sign up to the extended intervention (Get Healthy Service) and have at least one assessment with GHS (b) partially compliant if there is a record of GHS sign-up and initial assessment/engagement (c) fully compliant if they are noted as having graduated from the GHS program.

## 8 Statistical Analyses

### 8.1 Primary outcome Analysis

There are six primary hypotheses resulting from an all-pairwise comparison of the three study groups (two intervention groups and control) in the two primary outcomes of

1. maternal systolic blood pressure decrease
2. binary threshold on maternal weight (≥ 4kg reduction) and/or waist circumference (≥ 2cm reduction)

change from 6-12 months postpartum.

The three pairwise hypotheses relating to primary outcome 1 (BP) will be incorporated into an ANOVA model, and the three pairwise hypotheses relating to outcome 2 (composite measure) incorporated into a logistic regression model. The model fit statistics (*F-*test and χ^2^-test) from each of the models will be used to determine (with a threshold of α = 0.05) whether the relevant individual pairwise hypotheses should be evaluated. If the threshold is met, one of the multiple testing methods described in Section 6.5 will be applied. All (three or six) tests that pass to the second stage of testing will be considered to be one family of tests and tested as two-sided and based on a null hypothesis of no treatment effect. A multiplicity adjusted *p*-value of less than 0.05 will be considered significant in any of the tests for the primary outcomes.

Analyses will be adjusted for hospital site.

Subgroup analysis will occur as stipulated in Section 6.3.

### 8.2 Secondary outcomes Analyses

Analysis of the secondary outcomes will proceed in a similar manner to the primary outcomes, with tests for differences across groups performed as a first step using ANOVA testing for continuous outcomes and chi-squared test (from a logistic regression model) for categorical outcomes. For ordinal outcomes we will first do an ordinal regression to compare 12m and 6m categories within treatment groups, we will then compare groups. However, there will be a reduction in the required significance level to α = 0.01. Where there is evidence of differences amongst groups in the secondary outcomes, the same multiple testing method will be used to determine between which groups the differences exist, while controlling the familywise type I error rate at α = 0.01.

Analyses will be adjusted for hospital site.

Subgroup analysis will occur as stipulated in Section 6.3.

### 8.3 Secondary Analyses of primary and secondary endpoints

Use of the Get Healthy Service (GHS) will be analysed in a secondary analysis of both primary and secondary endpoints comparing women with GHS usage ≥ 4 phone calls versus no evidence of signing up to GHS. Analyses will be adjusted for randomisation group to assess whether outcomes are associated with GHS usage independently of the effect of the intervention.

## 9 Safety Analyses

### 9.1 Adverse Events

We don’t expect any adverse events for these interventions.

### 9.2 Clinical Laboratory Evaluations

Laboratories are accredited by NATA and use the same reference ranges. Evaluation is not needed.

## 10 Reporting Conventions

P-values ≥0.001 will be reported to 3 decimal places; p-values less than 0.001 will be reported as “<0.001”. The mean, standard deviation, and any other statistics other than quantiles, will be reported to one decimal place greater than the original data.

Quantiles, such as median, or minimum and maximum will use the same number of decimal places as the original data. Estimated parameters, not on the same scale as raw observations (e.g. regression coefficients) will be reported to 3 significant figures.

## 11 Quality Assurance of Statistical Programming

To provide high quality code that is understandable and allows reproduction of the analysis the following points will be followed:

Any outputs will have the

● date and time included
● the name of the code file that produced the analysis
● the author

At the start of any code file there will be a set of comments that give

● the author
● the date and time of writing
● references to inputs and outputs
● reference to any parent code file that runs the child code file

## 12 Summary of Changes to the Protocol

There are some minor changes to analyses compared to the original protocol:

- Addition of an alternative method to deal with multiple testing (see Section 6.5) to allow more flexibility about which statistical programs will be employed (R, SAS, Stata)
- Addition of hospital site as covariate for adjusting models and testing for interaction to account for differences in hospital practices (Section 6.3.1)
- Addition of Full analysis set secondary analyses, may help to provide more understanding about the effect of the intervention if the main analysis is null (Sections 6.2.2 and 6.2.3)

## Data Availability

All data produced in the present work are contained in the manuscript.

## Data Availability

All data produced in the present work are contained in the manuscript.

## Abbreviations and Definitions

BP2: Blood Pressure Postpartum Study
CH: Chronic hypertension
CH+PE: Chronic hypertension with superimposed preeclampsia
CRP: C-reactive protein
CVD: Cardiovascular disease
eGFR: Estimated glomerular filtration rate
GH: Gestational hypertension
GHS: Get Healthy Service
GP: General Practitioner
HDP: Hypertensive disorders of pregnancy
HOMA: Homeostatic assessment (of insulin resistance)
hsCRP: High-sensitivity C-reactive protein
PE: Preeclampsia
PP: Postpartum
SAP: Statistical Analysis Plan
SD: Standard Deviation
UNSW: University of New South Wales

## 14 Draft Tables and Figures

**Table 1.**
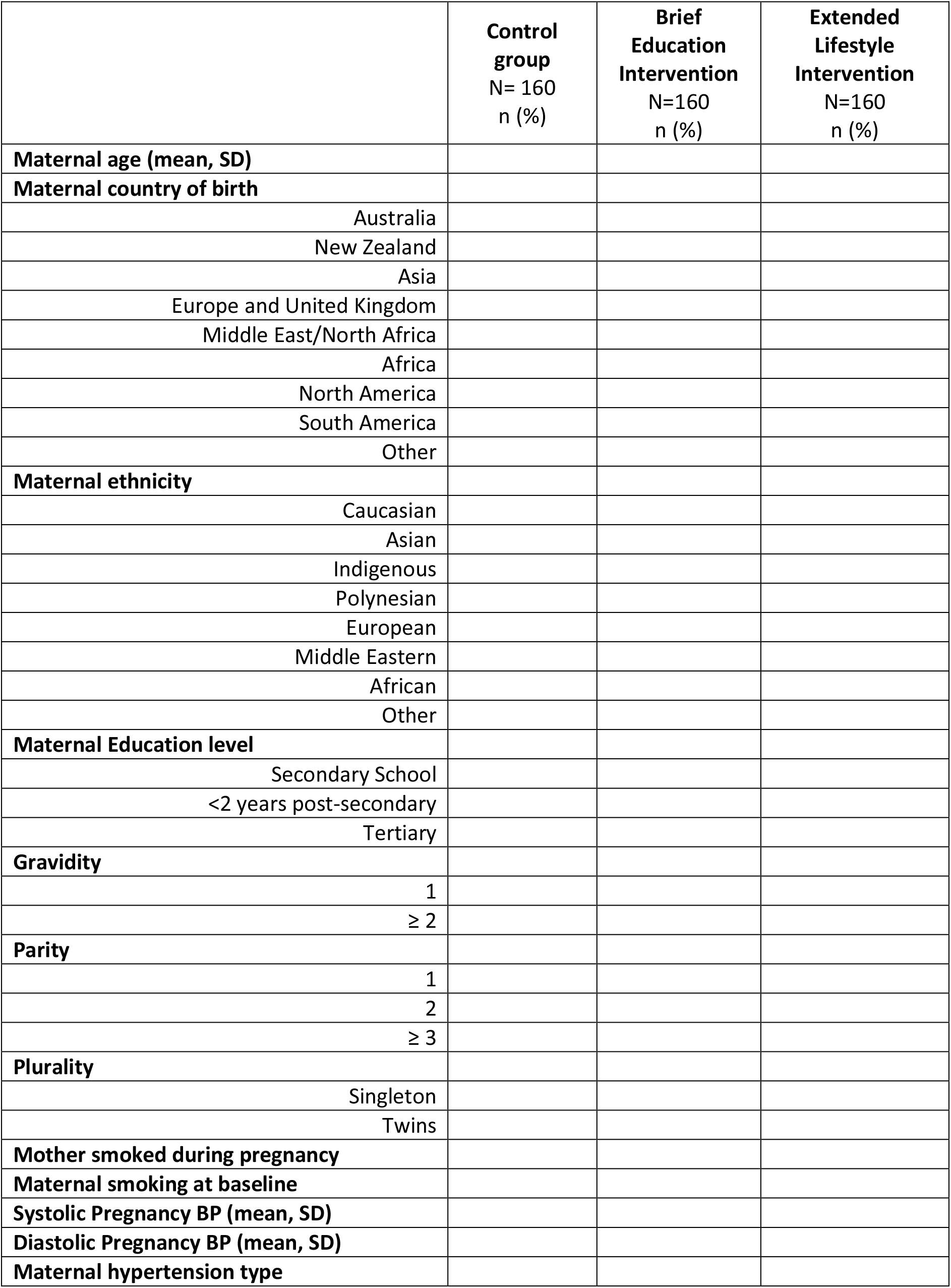
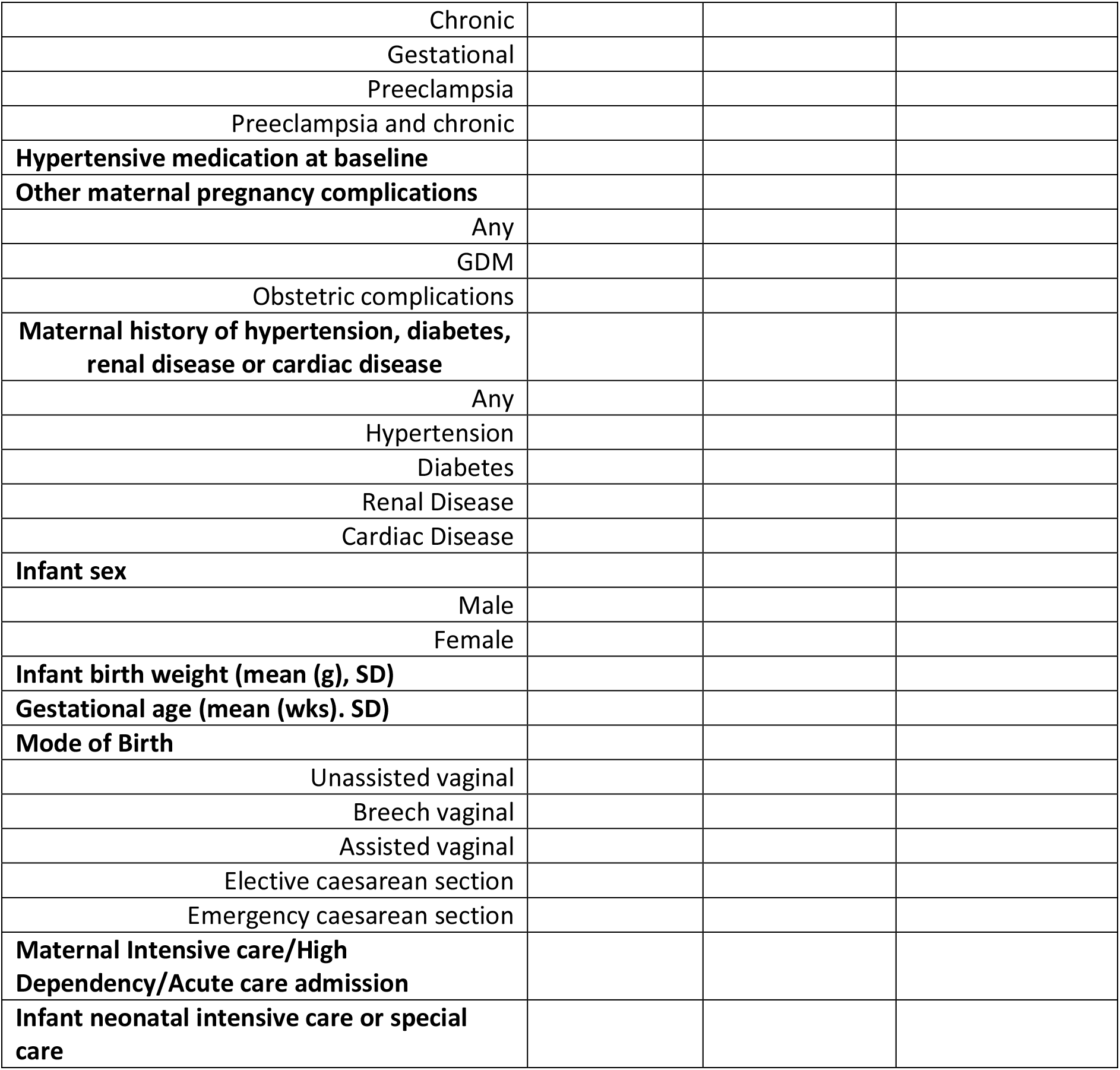
Descriptive characteristics of BP^2^ Study Population, index births and mothers.

**Table 2.**
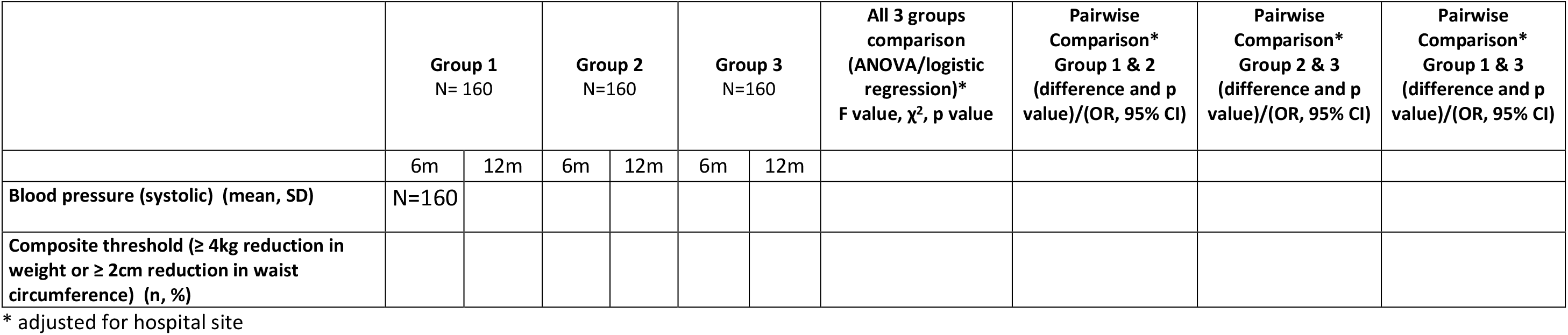
Primary outcome ITT analysis. Changes from baseline (6 months after birth) to end of intervention period (12 months after birth) in BP2.

**Table 3.**
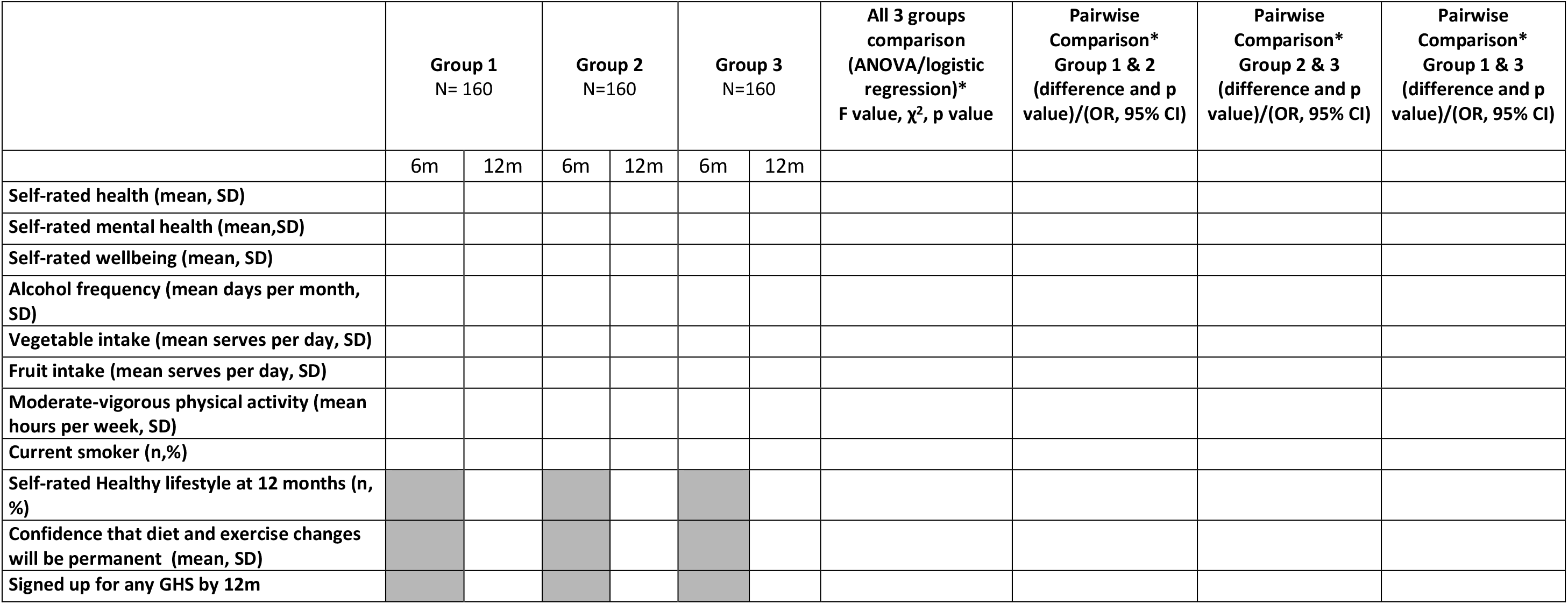
Secondary outcome analysis-Lifestyle Behaviour changes.

**Table 4.**
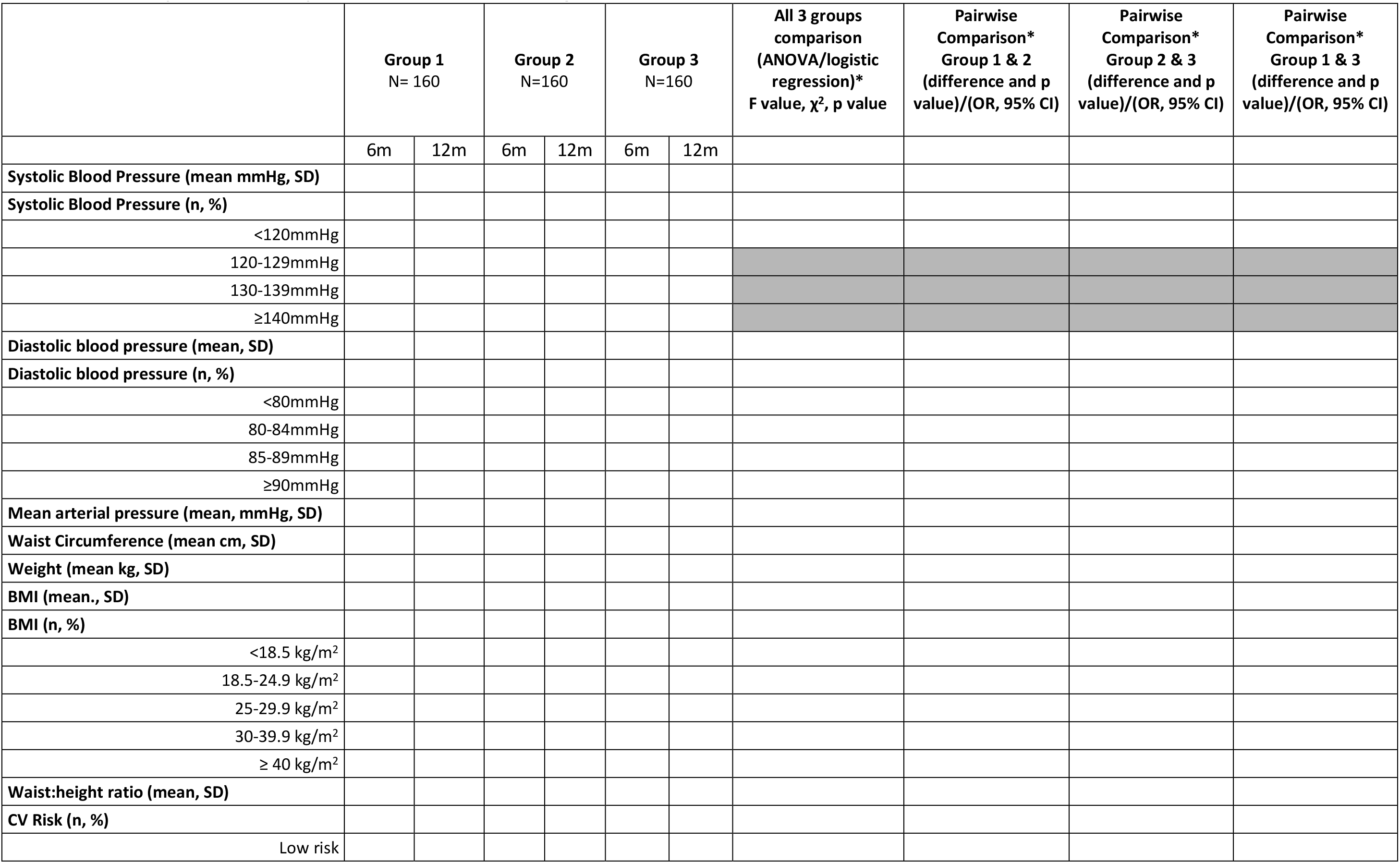
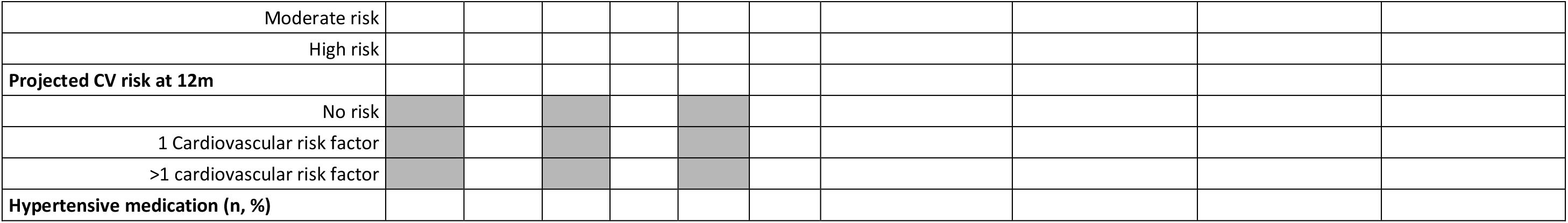
Secondary outcome analysis – Maternal Clinical changes.

**Table 5.**
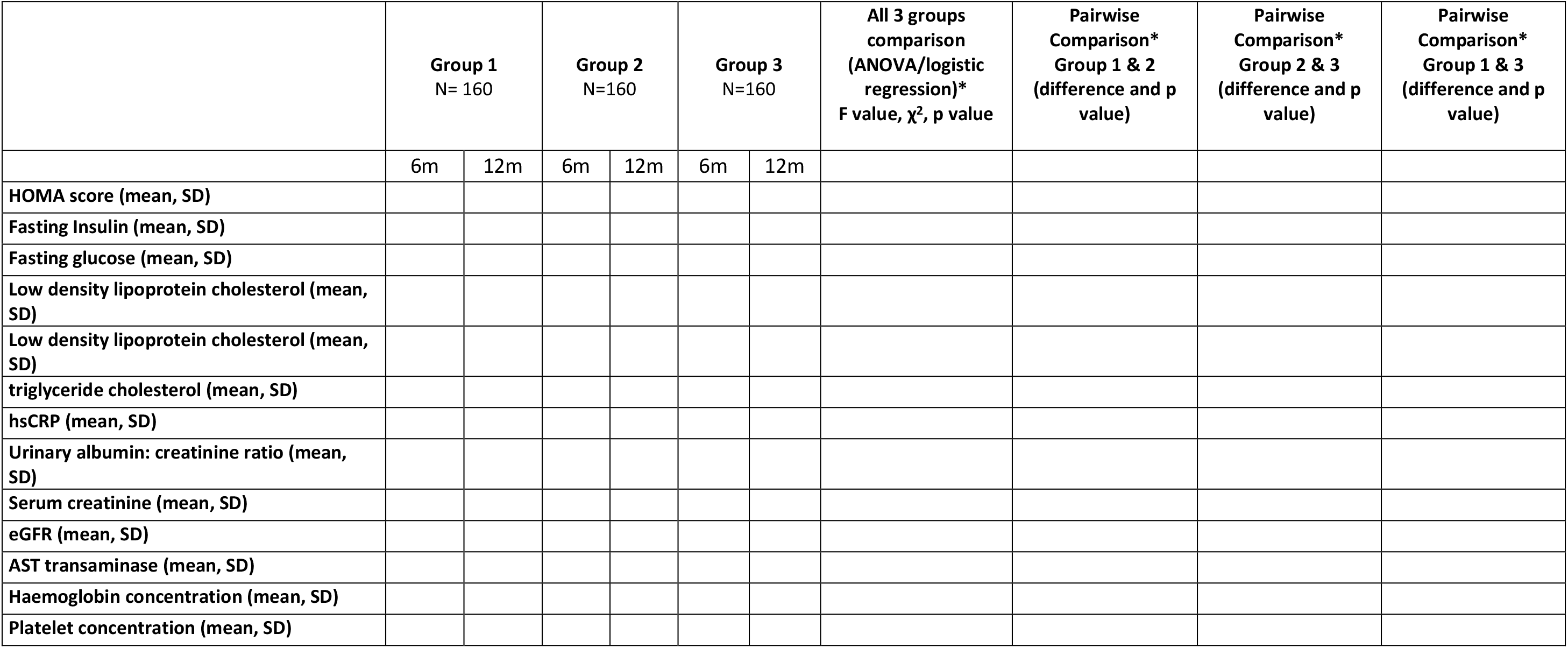
Secondary outcome analysis – Maternal Biochemical changes.

**Table 6.**
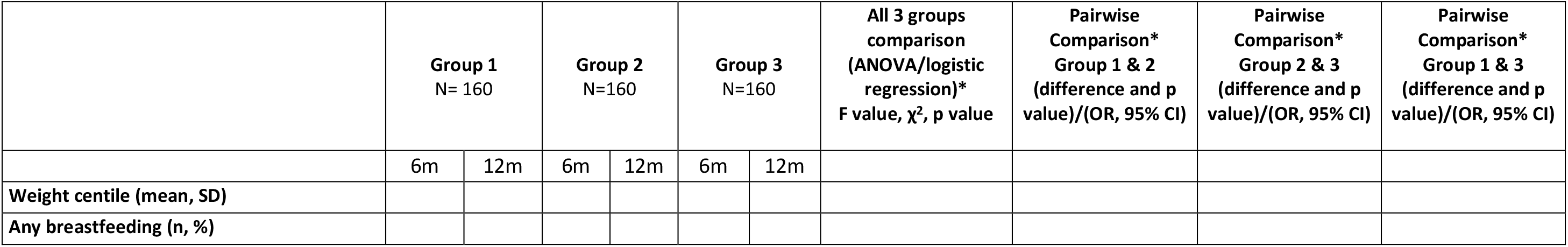
Secondary outcome analysis-Infant changes

